# Flexibility of brain dynamics is increased and predicts clinical impairment in Relapsing-Remitting but not in Secondary Progressive Multiple Sclerosis

**DOI:** 10.1101/2023.07.25.23293132

**Authors:** Lorenzo Cipriano, Roberta Minino, Marianna Liparoti, Arianna Polverino, Antonella Romano, Simona Bonavita, Viktor Jirsa, Giuseppe Sorrentino, Pierpaolo Sorrentino, Emahnuel Troisi Lopez

**Author notes:** **Correspondence to:** Prof. Pierpaolo Sorrentino, MD, PhD, Department of Biomedical Sciences, University of Sassari, Sassari, Italy. These authors contributed equally (Lorenzo Cipriano, Emahnuel Troisi Lopez).

## Abstract

**Background:** Large-scale brain activity has long been investigated under the erroneous assumption of stationarity. Nowadays, we know that resting state functional connectivity is characterised by aperiodic, scale-free bursts of activity (i.e. neuronal avalanches) that intermittently recruit different brain regions. These different patterns of activity represent a measure of brain flexibility, whose reduction has been found to predict clinical impairment in multiple neurodegenerative diseases such as Parkinson’s disease, Amyotrophic Lateral Sclerosis, and Alzheimer’s disease. Brain flexibility has been recently found increased in Multiple Sclerosis (MS) but its relationship with clinical disability remains elusive. Also, potential differences in brain dynamics according to the MS clinical phenotypes remain unexplored so far.

**Methods:** We studied brain flexibility through source-reconstruction of magnetoencephalography (MEG) signals in a cohort of 25 MS patients (10 RRMS and 15 SPMS) and 25 healthy controls (HC).

**Results:** RRMS patients showed greater brain flexibility than HC. On the contrary, no differences in brain dynamics were found in SPMS patients as compared to HC. Finally, brain dynamics showed a different predictive power on clinical disability according to the MS type.

**Conclusion:** For the first time, we investigated brain dynamics in MS patients through high temporal resolution techniques, unveiling differences in brain flexibility according to the MS phenotype and its relationship with clinical disability.

## 1. INTRODUCTION

Multiple Sclerosis (MS) is a chronic inflammatory disease of the central nervous system characterised by a complex association of both demyelination and diffuse neurodegeneration of the grey and white matter ^1^. Disease occurs most often with a clinical phenotype characterised by a relapsing remitting course (RRMS). However, about 50% of MS patients can evolve as a secondary progressive form (SPMS) and a minority may show worsening from the onset, the primary progressive form (PPMS) ^1,2^. Recent studies support the idea that RRMS and SPMS are part of a disease continuum in which the phase transition is driven by the change in the balance between inflammatory and neurodegenerative mechanisms ^3^. The different clinical presentations, which are a consequence of distinct underlying pathophysiologic mechanisms, may also explain the large variability of responses to the currently available immunosuppressive and immunomodulatory treatments ^1,3,4^.

Magnetic resonance imaging (MRI) is currently established as a key diagnostic tool in MS ^5^ due to its ability to detect the spatial and temporal distribution of MS-associated lesions. Nevertheless, years of use of MRI have shown that only a small fraction of MS clinical features and outcomes can be explained by the lesion load. This mismatch is called the clinico-radiological paradox and highlights our lack of understanding of this complex disease ^6^.

Many functional MRI (fMRI) studies deployed neural network theory in the attempt to shed light on the functional effects of the neuropathological processes that characterise the different clinical phenotypes. Taken together, these studies demonstrated relationships between changes in functional connectivity (FC) among specific brain regions and clinical features of the disease ^7–9^. However, these results were internally inconsistent, as they often failed to replicate. For instance, some studies found that increased resting state-FC (RS-FC) of selected brain regions/networks was related to better cognitive performances ^7,10^, while others showed an increased RS-FC of the same regions/networks in cognitively impaired individuals ^8,9^.

Potential explanations for these discrepancies include the temporal neuropathological evolution of the disease (i.e. different studies were performed in different disease stages), its clinical heterogeneity, the methodological differences across fMRI studies and, finally, the intrinsic limitations of the fMRI, including its low temporal resolution. The temporal smoothing induced by the slow haemodynamic response reduces the ability to evaluate fast brain reconfigurations at fast time-scales. Conversely, magneto- and encephalography (MEG, EEG) provide a more direct assessment of the brain’s fast activities ^11^.

Large-scale brain scans have been processed typically under the assumption of stationarity. However, now we know that RS-FC evolves over time in a non-linear fashion ^12^. In particular, brain activity is characterised by aperiodic, scale-free bursts of activity (i.e. neuronal avalanches) that intermittently interconnect brain regions ^13–15^ and that account for most of the time-averaged FC ^16^. In particular, aperiodic bursts reconfigure over time, giving rise to rich, non-stereotyped dynamics. In fact, healthy brains constantly recruit different brain regions generating a high number of patterns of activations. Thus, the number of such patterns represents a measure of brain flexibility dynamics, whose reduction has been found to predict clinical impairment in multiple neurodegenerative diseases ^17,18^.

Based on our previous observations about brain dynamics in neurodegenerative diseases such as probable Alzheimer’s disease (AD) ^19^, Parkinson’s disease (PD) ^17^ and Amyotrophic lateral sclerosis (ALS) ^18^, in the present work we hypothesise that MS could also be characterised by variations in brain flexibility, which would be related to, and predictive of, the subject specific clinical impairment. We also wondered whether the underlying disease mechanisms that characterise the two distinct forms of MS (relapsing remitting and progressive) could reflect in different brain dynamics and if this difference could help in predicting clinical disability.

To test these hypotheses, we source reconstructed MEG scans performed in twenty-five MS patients (10 RRMS and 15 SPMS) and twenty-five healthy controls. To estimate flexibility, we calculated the number of unique patterns of neuronal avalanches expressed in each MEG recording. Operationally, a neuronal avalanche is defined as an event starting when at least one brain region deviates from its baseline activity, and ending when all regions return to their normal level of activity. An avalanche pattern is defined as the set of all the brain areas that were recruited at any moment during an avalanche. The functional repertoire is defined as the set of the unique patterns that occurred over time and used its size as a surrogate marker of brain flexibility. Finally, we implemented a multilinear regression model with *k-fold* cross validation to verify the ability of the size of the functional repertoire to predict, at individual level, the clinical impairment assessed by the Expanded Disability Status Scale (EDSS).

## 2. MATERIALS AND METHODS

### 2.1. Participants

Twenty-five MS patients (five males and 20 females) and twenty-five age-, sex- and education-matched healthy controls (HC) were recruited. MS was diagnosed in accordance with the 2017 revision of the McDonald criteria ^5^. MS individuals were further classified in Relapsing-Remitting MS (RRMS) and Secondary-Progressive MS (SPMS). The eligibility of the patients was defined according to the following exclusion criteria: 1) use of illicit drugs, stimulants, amphetamines, barbiturates, and cannabis; 2) a history of central nervous system (CNS) disorder other than MS; 3) severe mental illness; 4) other systemic disorders with possible secondary involvement of the CNS.

MS patients underwent a clinical examination performed by an experienced neurologist. The EDSS ^20^ was used to evaluate disease-related disability. Fatigue was assessed by the Fatigue Severity Scale (FSS) ^21^ while neurocognitive function was evaluated by the Symbol Digit Modalities Test (SDMT) ^22^. Key symptoms of depression were studied through the Beck Depression Inventory (BDI) self-rated scale ^23^. The study protocol was approved by the Local Ethics Committee (University of Campania “Luigi Vanvitelli”) with protocol number 591/2018. All participants provided written informed consent in accordance with the Declaration of Helsinki.

### 2.2. MEG and MRI acquisition, preprocessing and source reconstruction

MEG acquisition, preprocessing, source reconstruction and connectivity estimation have been performed according to our previous studies ^24–27^. All the patients and healthy controls underwent magnetic resonance imaging (MRI) recorded by a 1.5-T GE Medical System (GE Healthcare, Milwaukee, MI, USA) scanner. Three-dimensional high-resolution T1-weighted (3D-T1) inversion recovery prepared fast spoiled gradient recalled sequence (IR-FSPGR, TR = 8.216 ms, T1 = 450 ms, TE = 3.08 ms, flip angle = 12, voxel size = 1 mm3 × 1 mm3 × 1.2 mm3) were acquired to extract volumetric data.

### 2.3. Analysis of brain dynamics

#### 2.3.1. Neuronal Avalanches and branching parameter

To quantify the spatio-temporal fluctuations of brain activity, we estimate neuronal avalanches. A single neuronal avalanche is defined as an event starting with a fluctuation of the regional brain activity, in at least one ROI, and ending with the return of all the involved ROIs to their normal activity ^28^.

Each of the source-reconstructed signals (derived from the 90 ROIs) was z-transformed and thresholded according to a cutoff of 3 standard deviations (i.e., z > |3|) ^17^. A confirmation of the results’ independence from the chosen threshold was performed by changing the threshold from 2.5 to 3.5.

To capture the critical dynamics, we binned the time series ^17^ by estimating the suitable time bin length by computing the branching ratio *σ* for each individual, for each avalanche and for each time bin duration ^13^. Specifically, the branching ratio was calculated as:

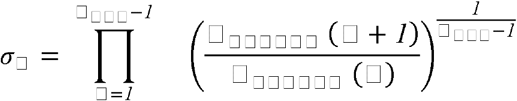

where σ is the branching parameter of the i-th avalanche in the dataset, Nbin is the total number of bins in the i-th avalanche, nevents j - is the total number of events in the j-th bin. After that we averaged the results over all avalanches as follow:

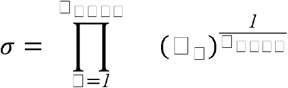

Critical processes are represented by σ = 1, that was present at bin length = 3. However, we repeated our analysis varying time bins from 1 to 5 and obtained similar results. Each avalanche had an avalanche pattern defined as the set of all ROIs that were above the threshold.

#### 2.3.2. Functional repertoire

For each individual, we calculated the functional repertoire as the number of unique avalanche patterns expressed during the recording ^17^. Unique indicates that each avalanche pattern is counted only once over the extent of the functional repertoire (i.e., repetitions are discarded).

### 2.4. Multilinear regression analysis

Starting from the assumption that fluctuations in brain dynamics could predict clinical impairment, we performed a multilinear regression model. The latter was performed by including a clinical feature (EDSS, FSS, SDMT or BDI) as a dependent variable and MS type and functional repertoire as independent variables. Multicollinearity was assessed through the variance inflation factor (VIF). To validate our approach, we performed *k*-fold cross-validation, with *k* = 5 ^29^. Specifically, *k* iterations were performed to train our model and at each iteration the *k*^*th*^ subgroup was used as a test set.

### 2.5. Statistical analysis

Statistical analysis was carried out in MATLAB 2021a and R Studio (http://www.rstudio.com/). A t-test and a Chi-square was used to compare patients and controls for age, educational level and sex. A Wilcoxon rank sum test was used to compare HC and MS groups. Kruskal-Wallis test was performed to compare HC, RRMS, and SPMS groups. The results were corrected by the false discovery rate, and the significance level was set at p-value < 0.05. The relationship between the size of the functional repertoire and the clinical scores was investigated in the MS group using the Spearman’s correlation coefficient. The predictive power of the flexibility parameter on clinical features has been investigated through a multilinear regression model.

## 3. RESULTS

### 3.1. Cohort characteristics

Sociodemographic and clinical characteristics of our cohort are reported in Table 1.

**Table 1.**
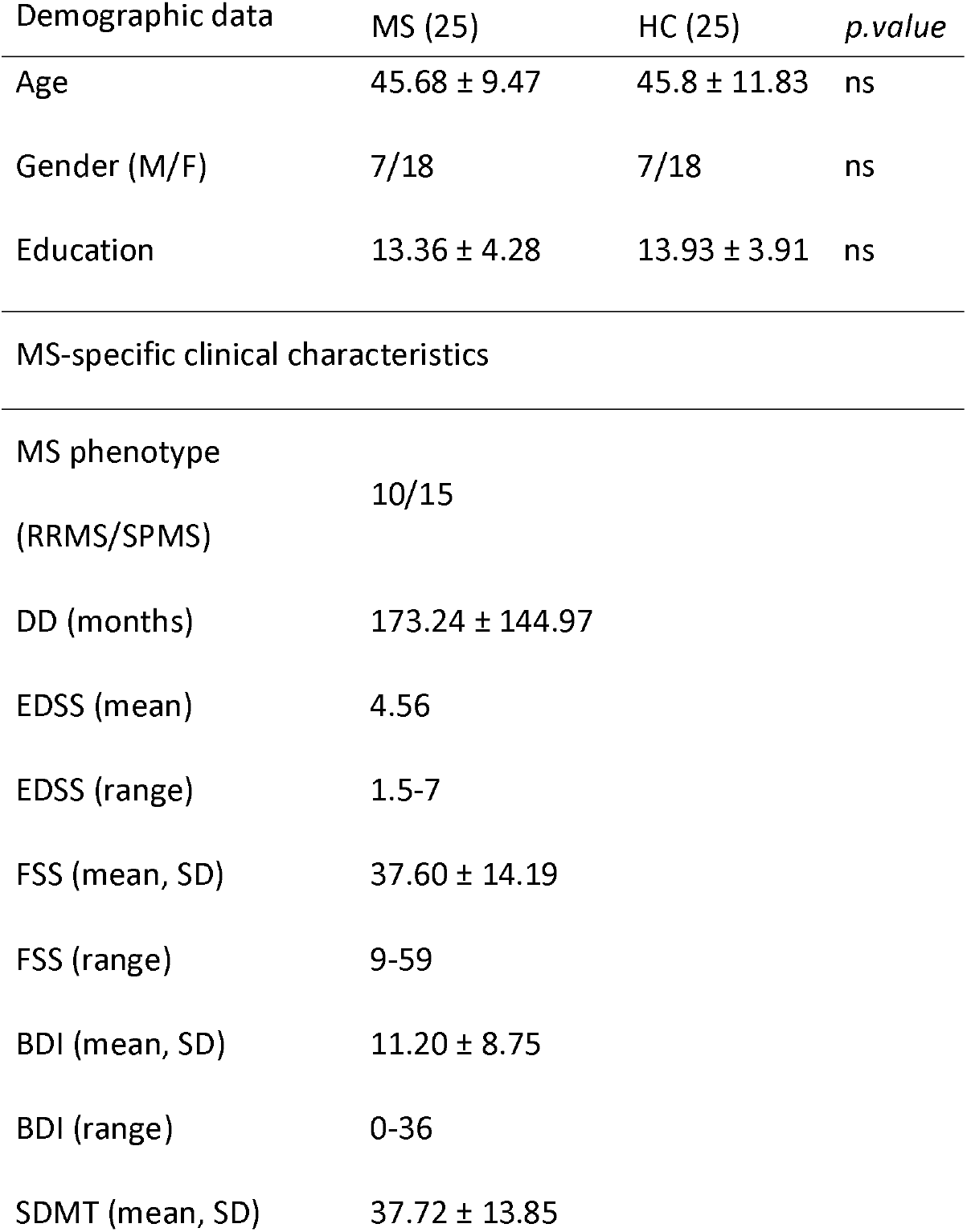
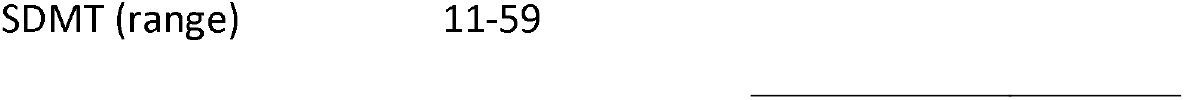
Sociodemographic and clinical characteristics of the cohort.

No significant difference in age, gender and education was found between the two groups. Abbreviations: MS: Multiple Sclerosis; HC: Healthy Controls; RRMS: Relapsing Remitting MS; SPMS: Secondary Progressive MS; DD: Disease Duration; EDSS: Expanded Disability Status Scale; FSS: Fatigue Severity Scale; BDI: Beck Depression Inventory; SDMT: Symbol Digit Modalities Test.

### 3.2. Analysis of brain dynamics: the functional repertoire

The comparison between MS and HC showed a larger functional repertoire in MS patients (p = 0.006) (**Fig. 2A**). We also performed a Kruskal-Wallis test to evaluate differences in the size of the functional repertoire according to the MS clinical form (χ2(2) = 9.8, p = 0.007) (RRMS and SPMS). As shown in **Figure 2B**, the difference in brain flexibility between MS and HC was primarily driven by the RRMS patients (p = 0.02).

**Figure 1:**
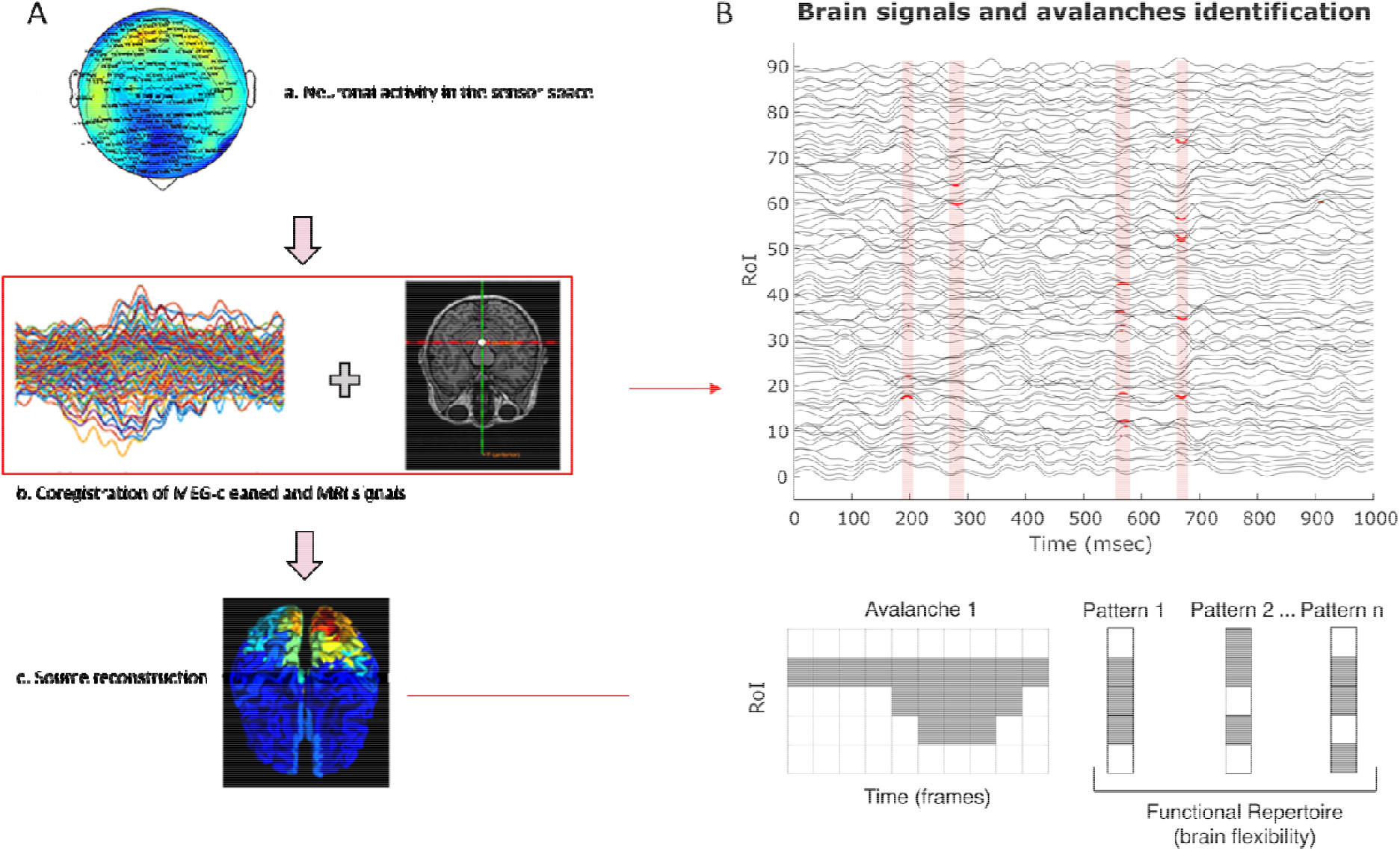
Pipeline overview and neuronal avalanches representation. Panel A: a) Registration of neuronal activity through magnetoencephalography (MEG); b) cleaned sensor signals (without physiological artefacts) co-registered with structural magnetic resonance imaging (MRI) of each participant. c) through a Beamformer algorithm, the time series of the sources were estimated in regions of interest (ROIs) within the brain according to a parcellation based on the Automated Anatomical Labelling (AAL) atlas. Panel B: the light red boxes represent the time frame in which a neuronal avalanche occurred. Specifically, the red dots indicate the frame in which the time series was above threshold (z-score > 3). In the bottom section of the panel, a schematic representation of a neuronal avalanche.

**Figure 2:**
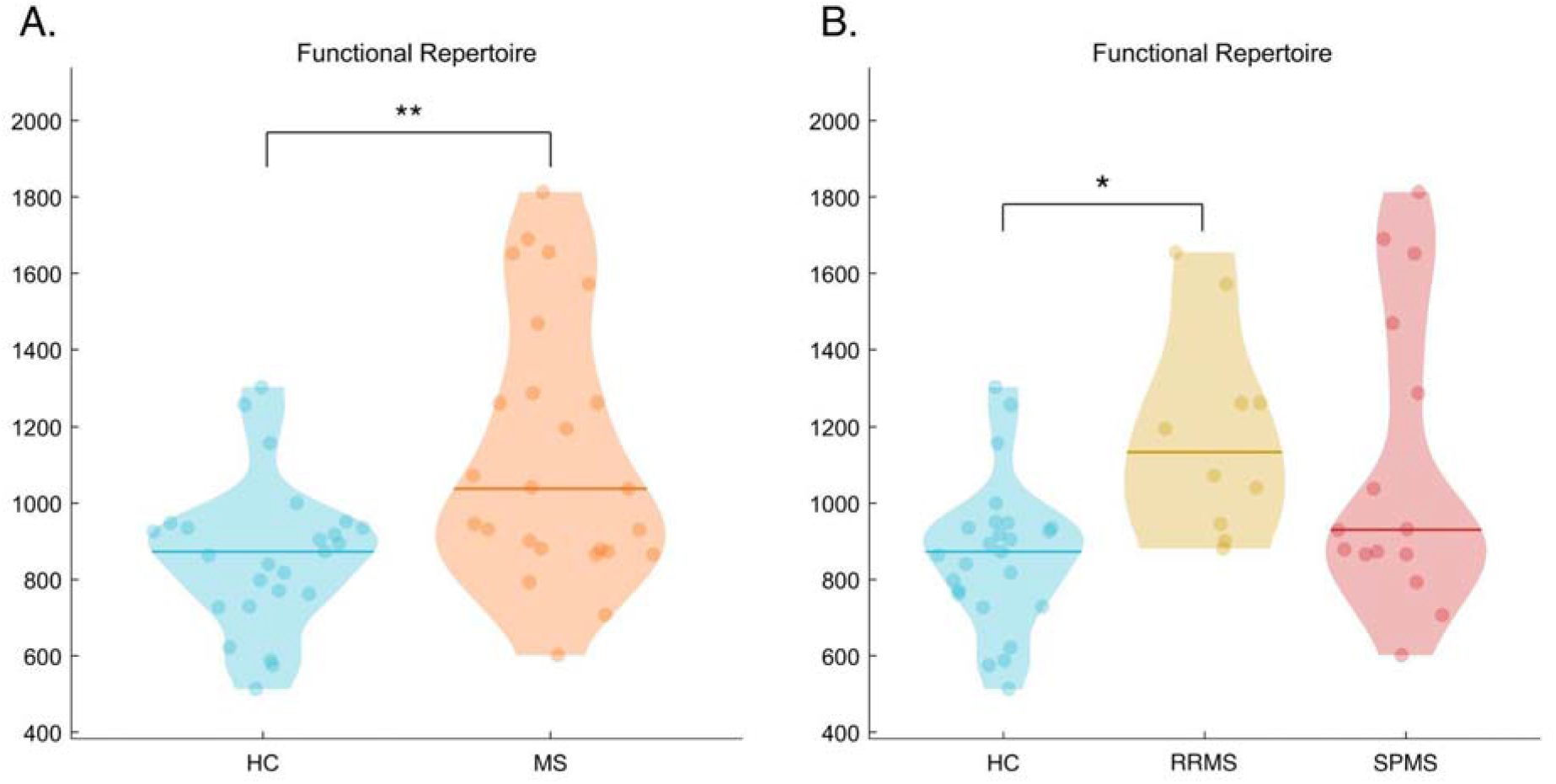
Brain flexibility comparison. **A**. Violin plots (healthy controls (HC) in blue and patients with multiple sclerosis (MS) in orange) of the number of unique avalanche patterns. **B. Functional repertoire according to the MS type**. Violin plots including MS type separation (Relapsing-Remitting Multiple Sclerosis (RRMS) in yellow, and Secondary Progressive Multiple Sclerosis (SPMS) in red. Significant p-values after false discovery rate correction: * p < 0.05, ** p < 0.01, *** p < 0.001.

### 3.3. Impairment prediction according to MS type

We performed a k-fold cross-validated multilinear regression analysis setting the clinical variables (FSS, EDSS, SDMT, BDI) as dependent variables, and tried to predict them by the means of the size of the functional repertoire. We also included the MS phenotype as further predictor and its interaction with the size of the functional repertoire to account for a possible different behaviour in the relationship between the functional repertoire and the EDSS score. We obtained a significant regression model (f (3,21) = 3.61, p = 0.03) able to predict 34% of the EDSS variance, with a normalised root mean squared error of the prediction equal to 25%. The significant contribution was determined by the MS type and its interaction with the size of the functional repertoire (beta coefficient = 0.461, p = 0.018 and beta coefficient = −0.456, p = 0.0419, respectively) (Fig. 3A). The interaction effect between MS type and functional repertoire suggests that the MS type is worthy in obtaining a significant prediction. Indeed, when removing the interaction effect, the model was not predictive anymore, while the MS type only, as a categorical variable, was not enough to predict the EDSS score. Given the MS type relevance, we observed the comparison between actual and predicted EDSS in each group separately (Fig. 3B). It can be noticed that while the predicted values of the RRMS group (yellow) showed a direct agreement with the actual values, this was not the case with the SPMS predictions (red). The distribution of the standardised residuals was then observed (figure 3C), and its absolute value was compared between RRMS and SPMS (Fig. 3D). The statistical comparison confirmed higher error in the prediction of the EDSS of the SPMS group (p = 0.017). We found no significant results when trying to predict FSS, SDMT and BDI.

**Figure 3:**
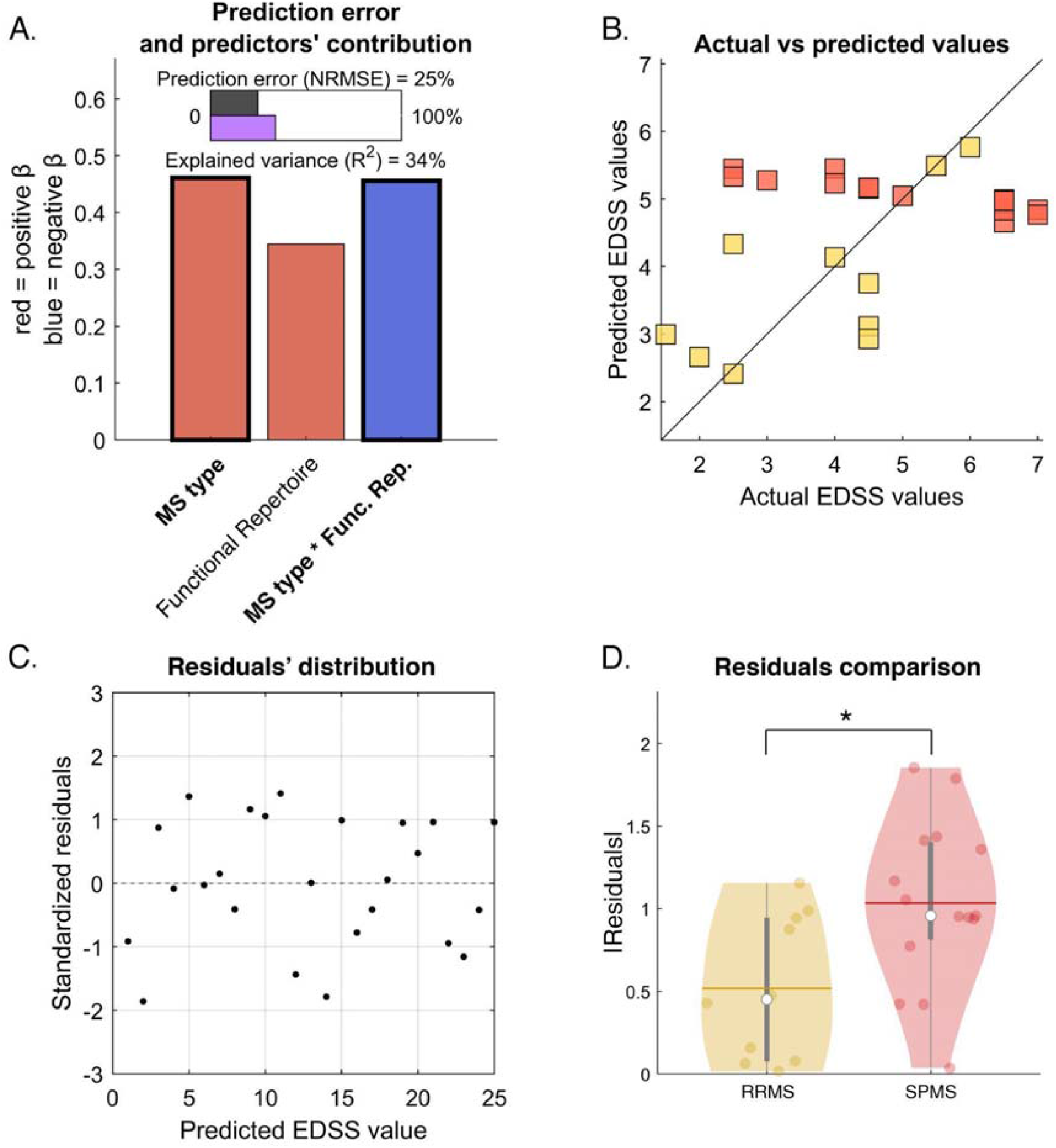
Clinical impairment prediction. Multilinear regression analysis with *k-fold* cross validation was performed to verify the ability of the size of the functional repertoire to predict clinical impairment assessed by the Expanded Disability Status Scale (EDSS). The clinical phenotype of multiple sclerosis (MS) was added as a predictor into the model (individually and as an interaction with the functional repertoire), to verify whether the prediction would depend upon the MS form (Relapsing Remitting MS (RRMS) or Secondary Progressive MS (SPMS). **A**. Regression analysis data: R2, normalised root mean square error (NRMSE); significant predictors in bold. **B**. Scatter plot to compare actual EDSS values with the EDSS values predicted through cross-validation. Since the MS type and the interaction between the MS type and functional repertoire were significant, we used different colours to represent RRMS (yellow) and SPMS (red) and observe the predictions independently. **C**. Scatter plot of the residual’s distribution. **D**. Statistical comparison between RRMS and SPMS absolute value of the residuals. Significant lower residuals indicate better prediction of the EDSS values of the RRMS group. * = p-value < 0.05.

The relationship highlighted by the regression model was confirmed through a Spearman correlation test, performed in each MS group, separately (Fig. 4). For SPMS patients no significant relationship was found between dynamical and clinical features (r = −0.13, p = 0.64). Conversely, in RRMS subjects the functional repertoire was significantly and positively related to the EDSS (r = - 0.7, p = 0.024). Similar results were evident also with the other clinical characteristics, but again, only in the RRMS group. In particular, SDMT and BDI showed respectively a significant negative (r = −0.64, p = 0.044) and positive (r = 0.65, p = 0.049) correlation with the size of the functional repertoire. A trend toward a positive statistically significant correlation was found for the fatigue scale. No significant (or nearly significant) relationships between clinical data and brain flexibility was found in the SPMS group.

**Figure 4:**
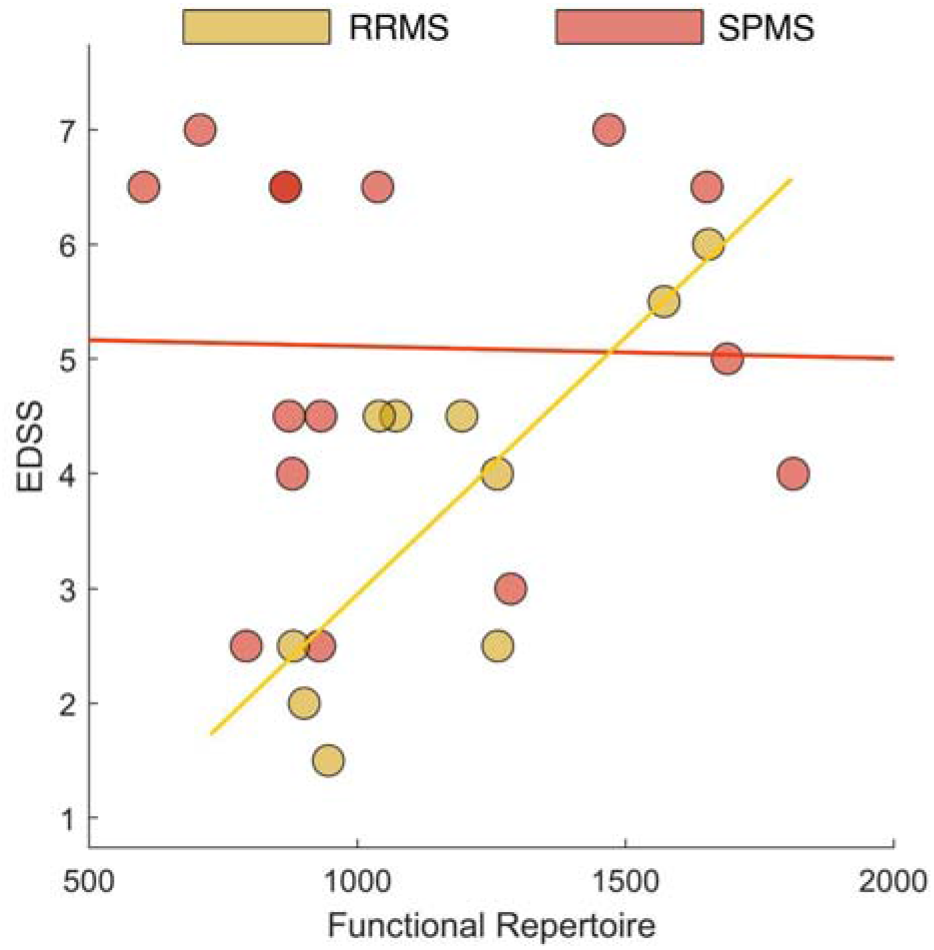
Correlation between EDSS and brain flexibility. Spearman correlation test between Expanded Disability Status Scale (EDSS) and functional repertoire was performed in Relapsing Remitting Multiple Sclerosis (RRMS) and Secondary Progressive Multiple Sclerosis (SPMS), separately. Significant correlation was found in the RRMS group only.

We also searched for a potential relationship between disease duration and brain flexibility in SPMS, performing a Spearman’s correlation that showed a trend toward a negative correlation between brain dynamics and disease duration (r = −0.48, p = 0.067).

## 4. DISCUSSION

In the present study, we investigated the flexibility of the brain dynamics in MS and its relationship with the clinical phenotypes. In particular, following our previous studies carried out on neurodegenerative diseases, including ALS ^18^, AD ^19^ and PD ^17^, where a reduction of the functional repertoire has been consistently found, we hypothesised that MS could express different behaviours as a function of the prevalence of neuroinflammation or neurodegeneration. When we looked at the different MS clinical phenotypes, we found that the significant differences in the flexibility of the brain dynamics were mainly attributable to the RRMS phenotype. In particular, RRMS patients showed a large number of unique patterns when compared to HC, whereas the dynamics in SPMS patients did not differ significantly as compared to HC.

In the second part of our work, we searched for relationships between flexibility and MS clinical features. Even in this case, RRMS and SPMS behaved in two opposite ways. The size of the functional repertoire did not show significant predictive power on EDSS. Nevertheless, the interaction between the size of the functional repertoire and MS phenotype showed significant predictive power. That is to say that brain dynamics show different predictive power according to the MS phenotype. In particular, brain dynamics positively correlated with disability in RRMS patients without showing significant relationship in SPMS patients. This means that only for the RRMS the increased number of patterns was associated with a worse clinical condition. Accordingly, in the RRMS group, a significant correlation between increased brain flexibility and other clinical signs such as depression and cognitive impairment (assessed by BDI and SDMT, respectively) was also found.

In our opinion, these results could hide two different explanations. On one hand the observation that only RRMS patients showed larger functional repertoires as compared to HC (and not SPMS patients) could suggest that the increased number of reconfigurations observed in RRMS brain might represents a compensatory mechanism adopted by the nervous system in the early phase of disease to maintain a proper functionality. On the other hand, the difference in brain dynamics between RRMS and HC could be due, at least partially, to the different pathophysiology that characterises RRMS and SPMS.

However, the results on the correlation between brain flexibility and clinical condition in RRMS subjects would seem to reject the compensatory hypothesis. In fact, when we performed statistical correlations between brain dynamics and clinical characteristics such as processing speed, depression, fatigue and disability, we found a positive relation between higher brain flexibility and worse clinical condition. All this seems to make it less probable a compensatory mechanism, except if the compensatory mechanism is not efficient. On the contrary, the neuropathological differences between the two MS phenotypes could partly explain the diversity in brain flexibility according to the clinical form of the disease. In fact, RRMS is a phenotype with predominant neuroinflammation and demyelination, whereas neurodegeneration, independent from the inflammatory responses, represents the main mechanism of SPMS disease progression ^3,30^. Hence, while the RRMS pathology is dominated by a peripheral immune response (brain parenchymal lymphocytes infiltration through a disrupted blood–brain barrier) that leads to the formation of new active lesions ^31^, progressive MS is characterised by subpial demyelinated lesions, with slow expansion of pre-existing white matter lesions and, most importantly, diffuse grey matter neurodegeneration ^1^. Speculatively, one can suppose that the coexistence of neuroinflammation and neurodegeneration could result in an unchanged level of overall flexibility in SPMS patients. In other words, the similar overall flexibility observed in SPMS with respect to HC may be the result of two concurrent mechanisms that affect the brain dynamics in opposite ways. One pattern would lead to more stereotyped brain dynamics, as previously found in neurodegenerative diseases. A different effect, as seen in the predominantly focal inflammatory RRMS phenotype, would lead to increased heterogeneity of the repertoires. In other words, the disruption of the myelin sheath observed in RRMS would lead to a dysregulation of the overall dynamics, which would become less effectively controlled, resulting in a higher number of states. Widespread degeneration, on the other hand, prevents the brain from accessing certain configurations, which would result in an impoverished repertoire. A comparable amount of both neurodegenerative and focal neuroinflammatory processes might contrast each other ^28^. According to this line of thought, we found a trend towards an inverse correlation between the disease duration and the flexibility of the brain dynamics in SPMS patients. That is to say, the more SPMS patients are close to the early phase of the disease (and, thus, the more similar they are to RRMS), the greater the flexibility of the brain dynamics, while longer disease durations (with increased neurodegenerative load), correspond to impaired flexibility.

Overall, our results are in line with the very recent studies that, through different neuroimaging techniques, investigated brain dynamics in MS. von Schwanenflug et al. ^32^ conducted a fMRI study in a cohort of almost entirely RRMS subjects, showing an increased flexibility of brain dynamics in MS patients. Similarly, by means of fMRI, Broaders et al. evaluated the brain dynamics in a cohort consisting of around 80% RRMS subjects. They found a greater number of brain network reconfigurations in patients with higher cognitive impairment ^33^. These results are in agreement with our findings that show a positive relationship between worse clinical condition and higher brain flexibility in RRMS subjects.

Some limitations of the current work should be pointed out. Firstly, the small sample size of the studied population. A second limitation is the absence of CSF/serum inflammatory biomarkers (or neuroinflammatory PET imaging biomarkers) to support the primary role of inflammation in affecting brain dynamics. An adjunctive weakness is the absence of a follow-up of the RRMS patients to demonstrate a change of brain dynamics in case of disease conversion to the progressive form. Future longitudinal studies evaluating the brain flexibility changes across the disease progression (and conversion) will be mandatory to confirm our hypothesis.

To our knowledge, we are the first to investigate brain dynamics through high temporal resolution techniques (M/EEG) in both RRMS and SPMS patients. These findings support the key role of temporal dynamics in understanding the link between brain connectivity and clinical features, also unveiling different dynamical features according to the MS phenotype. Additionally, if the supposed link between brain dynamics, inflammation and clinical outcome will be confirmed, this could allow monitoring in a minimal invasive and objective way (M/EEG analyses) the efficacy of MS immunotherapy ^4,34,35^.

## Declarations

## Author contributions

Concept and design: LC, ETL, PS, GS. Data collection, analysis and interpretation: RM, ML, AR, ETL, SB, VJ, PS, LC. Drafting of the manuscript: LC and ETL. Critical revision of the manuscript for important intellectual content: VJ, SB, GS.

## Data availability

The data that support the findings of this study are available from the corresponding author, GS, upon reasonable request.

## Ethical statement

All procedures performed were in accordance with the ethical standards of the institutional research committee and with the ethical standards laid down in the 1964 Declaration of Helsinki and its later amendments.

## Informed consent and consent to participate

Written informed consent has been obtained from all participants.

